# COVID-19 Associated Hepatitis in Children (CAH-C) during the second wave of SARS-CoV-2 infections in Central India: Is it a complication or transient phenomenon

**DOI:** 10.1101/2021.07.23.21260716

**Authors:** Radha Kanta Ratho, Ajit Anand Asati, Nitu Mishra, Ashish Jain, Sumit Kumar Rawat

## Abstract

**Objective:** While pediatric population has largely remained free of severe COVID-19, in some cases SARS-CoV-2 infection has been associated with complications like Multiple Inflammatory Syndrome in children (MIS-C). We mention another unique presentation subsequent to asymptomatic infection of SARS-CoV-2, a unique form of hepatitis designated by us as COVID-19 Associated Hepatitis in Children (CAH-C). The contrasting clinical presentations, temporal association and viral parameters of CAH-C cases, to the MIS-C cases are presented here.

**Methods:** As a retrospective and follow-up observational study we reviewed all children testing positive for SARS-CoV-2 during study period. Children presenting with “sudden onset of hepatitis, elevated transaminases, non-obstructive jaundice, lacking marked inflammatory responses and without evidence of (a) other known causes of acute hepatitis or previous underlying liver disease (b) multi-system involvement” were classified as CAH-C, are described here.

**Results:** Among 475 children tested positive, 47 patients presented with hepatitis, 37 patients had features of CAH-C, having symptoms of hepatitis only, with un-elevated inflammatory markers and uneventful recovery following supportive treatment. Whereas remaining 10 MIS-C hepatitis had protracted illness, multiple system involvement, required admission to critical care, and had mortality of 30%.

**Conclusion:** With the emergence of newer variants of concern (VOC) including the Delta variant which predominated the second wave of infections in India and has now spread to more than 142 countries with changing presentations, CAH-C might be one of them. Cases of such new entities need to be identified early and differentiated from other emerging syndromes in children during the ongoing pandemic for preventing adversities by timely intervention.

**Conflicts of interest:** The authors declare that they have no conflicts of interest related to the study or its findings. All authors have contributed to the conceptualization and manuscript writing of the study, the final version is approved by all the authors. We declare there are no competing interests involved among the authors.

**Funding and ethics approval:** Current research received no specific grant from any funding agency in the public, commercial or not-for-profit sectors. The follow-up and analysis work was performed after obtaining due approval of human ethics committee of the institution (Ref no. IEC/BMC/80/21).

## Introduction

Severe acute respiratory syndrome coronavirus 2 (SARS-CoV-2), the begetter of novel COVID-19 pandemic, was first identified in China in December 2019, with its subsequent epicenter being recognized in West, thereafter reached India causing the first wave(1). Interestingly during Initial phase of the pandemic children were disproportionately spared from severe illness with predominance of asymptomatic or mild cases, obviating need for hospitalizations(2,3). Subsequently, the “multisystem inflammatory syndrome in children” (MIS-C), which was first observed in children and adolescents in UK in April 2020, raised alarms about their safety more so in the absence of vaccines approved for children (4,5). This was soon followed by the detailed reports and linkage of MIS-C to SARS-CoV-2(6).

The initial phase of pandemic caused numerous infections among children in India but no serious illness(7). However, during the massive second wave of COVID-19 by April 2021 the cases involving children with severe illnesses increased(8). Newer variants of concern (VOC) including the delta (B.1.617.2) variant which spread to more than 142 countries from here, was largely responsible for this massive upsurge(9). This sometimes resulted in serious disease presentations varying from respiratory disease, to entities like MIS-C, encephalitis and acute hepatitis in children(10,11).

Although initial reports did not mention much about hepatitis as a one of the features or associated complications of COVID-19 (12), this finding was recognized in the later part of the year 2020. It has been observed in a subset of obese children with MIS-C, that severe disease and adverse outcomes were associated with co-existing hepatitis, similar to the findings observed among adults with COVID-19(13). However, about 60% of children with MIS-C presented with hepatitis in contrast to only 20-30% adult cases admitted with COVID-19, which has been linked to severe disease course among such patients(14,15). These findings indicate that age-related differences might exist in cases of hepatitis and the associated disease phenotypes. Moreover, its interplay with host immune response might have a bearing on the resulting morbidity, its management strategies, disease burden, and outcomes(16). During active COVID-19 infection, milder disease phenotype was observed without necessitating hospitalizations whereas more severe patterns were seen in cases with MIS-C often associated with prolonged need for hospital admissions. During the ongoing phase of the pandemic, apart from MIS-C there appeared a group of children presenting with hepatitis. Among these children such unique presentation of hepatitis, temporally associated with SARS-COV-2 infections is designated by us as CAH-C in addition to the cases of MIS-C. Such hepatitis cases outnumbered MIS-C cases and lacked the hallmarks of inflammation seen in MIS-C. This form of transient hepatitis in CAH-C is different from already familiar liver injury phenotypes and occasionally witnessed persistent disease in children having COVID-19.

In the current context, the cases of severe pediatric hepatitis of unknown origin affecting more than 169 children in nearly 12 countries, with at least 16 children had required liver transplantation(17). The effects of liver injury are unexplained by either SARS-COV-2 or Adenovirus alone in the absence of common hepatitis causing agents like hepatitis A-E(18). The underlying mechanisms, the role of a co-factor in play, if any and the respective dominant hepatic phenotype in such cases may have a link with emerging VOCs including the Delta, Omicron or further variants(19). Early identification of such entities is critical for early response, resource allocation, and formulating the guidelines for appropriate management. Our current study was planned with aim to correctly identify such cases, find out its temporal relation with SARS-CoV-2 infection and differentiate it from the other similar entities.

## Methods

### Study Design

It was a retrospective and follow-up observational study of record review from April 2021 to July 2021 at a tertiary care public hospital central India, which is a dedicated COVID-19 (500 beds) center for the entire region. Among persons tested COVID-19 positive by Indian Council of Medical research (ICMR) recommended RT-PCR assay, children presented with serious symptoms including hepatitis, were admitted to the hospital, the study included record review for all such cases along-with follow-up information as shown in *figure 1*. As per the existing protocols, many of these were referred through central control command telemedicine center from the districts smart city control room(20)

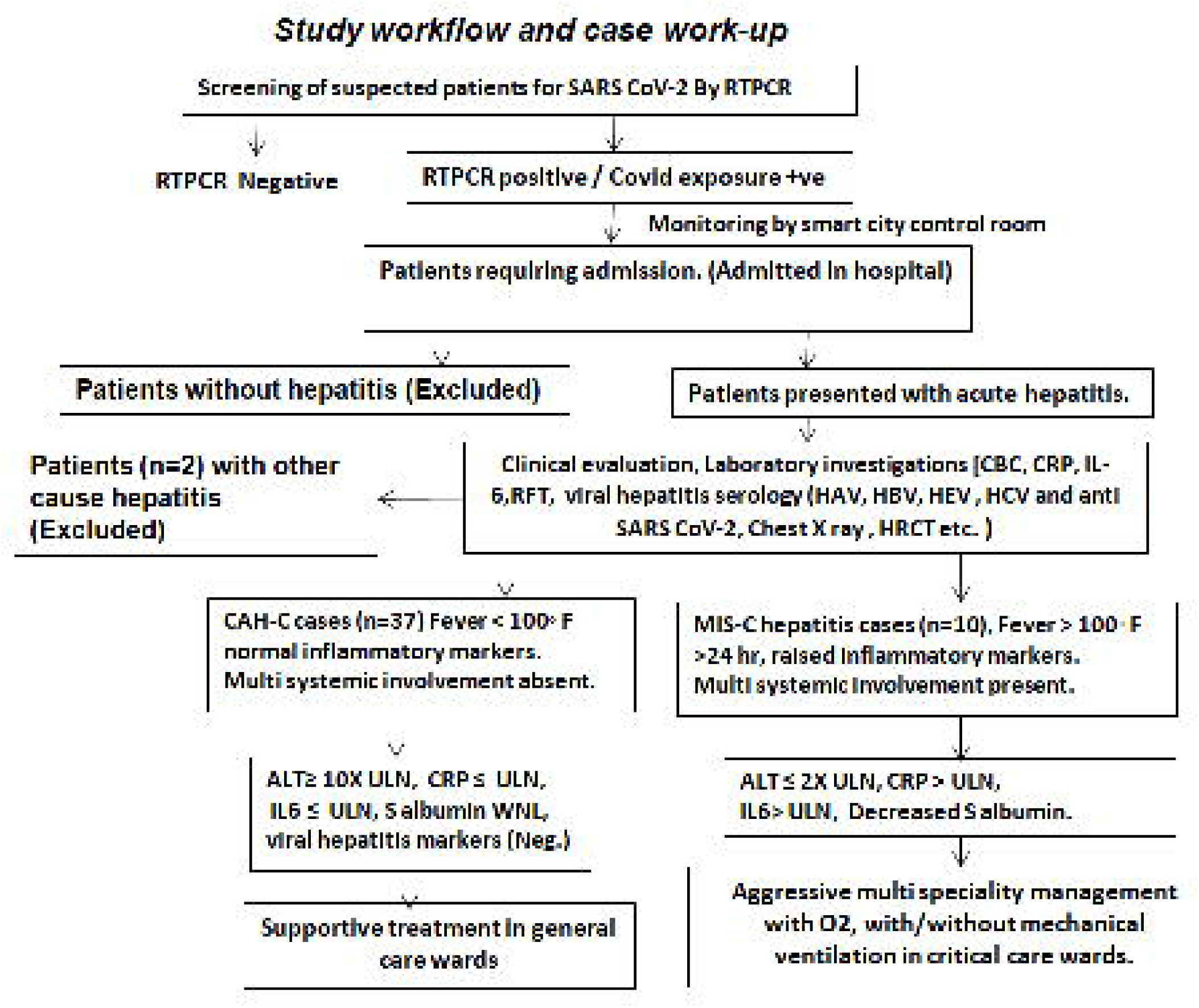

#### Inclusion criteria

Children within 14 years of age with laboratory evidence of COVID-19 during the study period.

Cases presented with hepatitis were categorized into two entities considering the temporal association in relation to COVID-19, disease severity (admission in intensive care unit), inflammatory marker levels and presence/absence of multi-system involvement (based on clinical, laboratory and radiological findings):

1. CAH-C: Those aged within 14 years, with a laboratory evidence of recent COVID-19 presenting with “sudden onset of hepatitis, elevated transaminases, non-obstructive jaundice, lacking marked inflammatory responses and without evidence of (a) other known causes of acute hepatitis or previous underlying liver disease (b) multi-system involvement”.
2. MIS-C associated hepatitis: Cases of MIS-C within definitions according to the Centers for Disease Control and Prevention (CDC), presenting along-with acute hepatitis i.e. elevated transaminases(21).

#### Exclusion criteria

Those with evidence of pre-existing liver disease, drug-induced liver damage or known cause of acute hepatitis were excluded.

### Laboratory investigations

SARS-CoV-2 RT-PCR screening tests were performed at the ICMR recognized Virology Research and Diagnostic Laboratory (VRDL) of the hospital. All such positive cases and COVID-19 suspects presenting with hepatitis were thoroughly evaluated as per the standard protocols including complete blood counts, liver function tests, renal function tests, serum ferritin, CRP, D-Dimer, LDH, Interleukin-6 (IL-6), myocardial enzymes, and procalcitoninin the Central clinical lab of the hospital. These children were retested for SARS-CoV-2 RT-PCR and further evaluated with additional tests including Adenovirus RT-PCR, HHV 1&2 RT-PCR, HbsAg, anti-HAV IgM, anti-HEV IgM, anti-HCV antibody, anti-Leptospira IgM, anti-EBV IgM, anti-VZV IgM, Widal test, malaria antigen, malaria antibody, antinuclear antibody (ANA), anti LKM antibody, total IgG, anti-SARS-CoV-2 IgG, dengue NS1 antigen, dengue IgM using ICMR recommended ELISA kits(22).

### Outcomes

Status at 4 weeks to discharge was taken as an endpoint for the assessment of outcomes.

### Statistical analysis

The data was extracted and entered in MS excel, proportions and percentages were calculated for categorical variables. The chi-square test was performed to find possible associations between the categorical variables like association of male sex, clinical outcome with either of entities. After checking the normality of data, *t*-test was applied to compare the mean values of different parameters. For those variables not having normality Mann Whitney U test or Fischer’s exact test was applied; A *p*-value of less than 0.05 was considered significant. The statistical analysis was performed using SPSS trial version 16 for windows.

## Funding and ethics approval

Current research received no specific grant from any funding agency in the public, commercial or not-for-profit sectors. The follow-up and analysis work was performed after obtaining due approval of human ethics committee of the institution (Ref no. IEC/BMC/80/21).

## Results

During the study period from April 2021 to July 2021 among the screened population of 15873, 475 (2.99%) children with a male to female ratio of 303:172(1.76:1) were found to be COVID-19 RT-PCR positive. The age group of these children ranged from 4 months to 14 years with mean age of 9 ± 3 years as shown in *figure 2. A*mong the admitted patients, 47 presented with features of hepatitis could satisfy the inclusion criteria, 2 cases presenting with hepatitis were excluded from the study as one of them was suspected to have drug induced liver injury, other was positive for HAV infection (*figure 1)*.As per the above-mentioned criteria, 37 patients (Male: Female, 23:14) and 10 (Male: Female, 3:7) patients could be categorized into CAH-C and MIS-C respectively *(Table 1)*. Majority of these children belonged to 2-6 years’ age group (*Figure 2*). There were no differences in their racial characteristics.

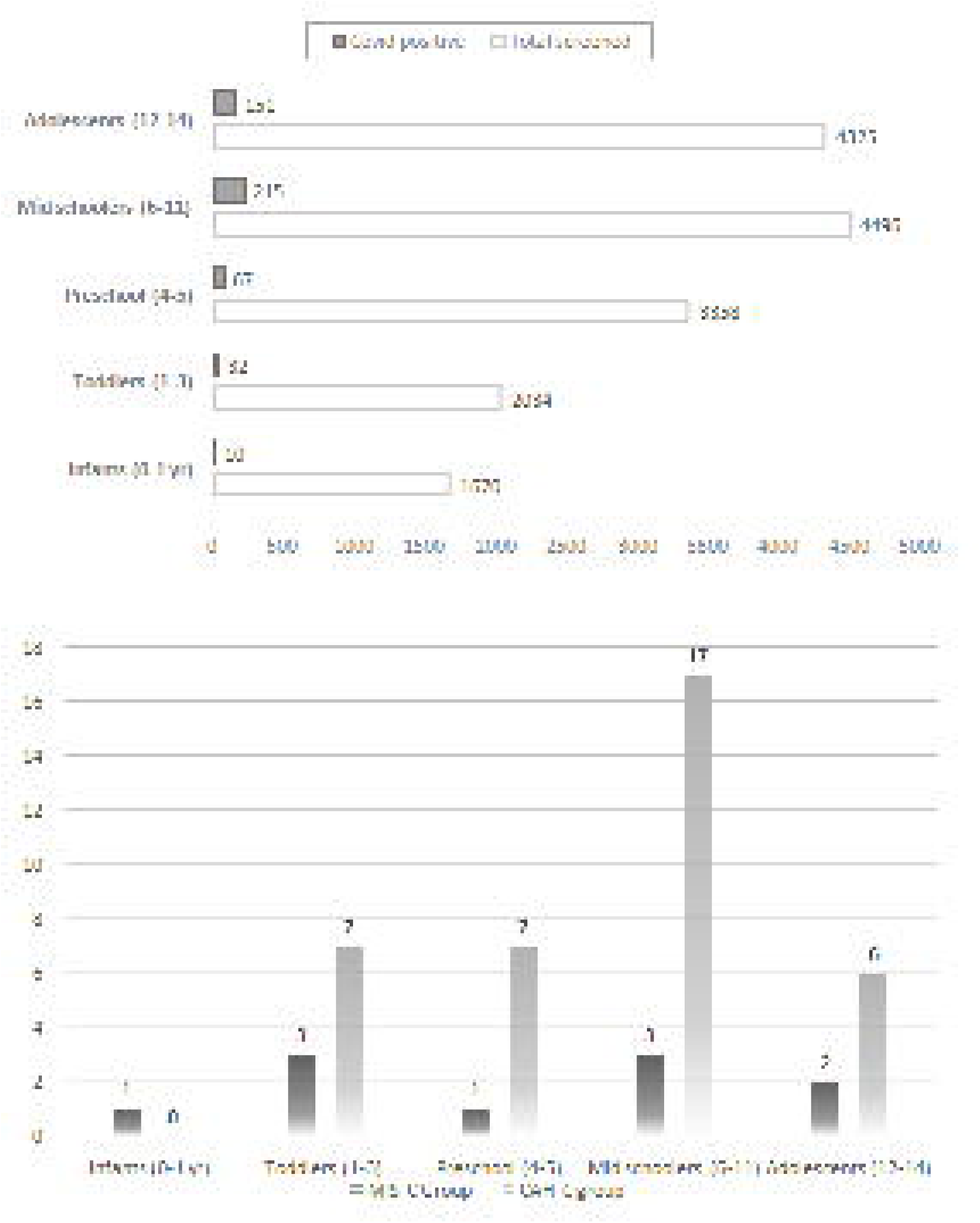

**Table 1:** Patient characteristics of CAH-C Cases Vs MIS-C associated hepatitis cases.

| Patient characteristics | CAH-C (n=37) | MIS-C (n=10) | P-value*<sup>\$</sup> |
| --- | --- | --- | --- |
| Age, mean ± SD, years | 6.75 (3.32) | 6.2 (4.29) | 0.3367 * |
| Male sex, No. (%) | 23 (63.9%) | 3 (30%) | 0.077 <sup>\$</sup> |
| Total days admitted, mean ± SD | 4.64 (1.37) | 8.4 (1.95) | 0.0001 * |
| Day from contact history, mean ± SD | 23.56 (6.63) | 9.7 (93.74) | 0.0001 * |
| Clinical outcomes: |  |  |  |
| Recovered, n (%) | 37 (100%) | 7 (70%) | 0.013 <sup>\$</sup> |
| Death, n (%) | - (-) | 3 (30%) | 0.013 <sup>\$</sup> |
| *Independent samples t-test, <sup>\$</sup> Fischer's exact test. | | | |

### Temporal relationship

The cases of CAH-C started appearing in April-end and peaked in may-end and then showed a decline in trend with small number of cases still presenting after 4 weeks of disappearance of new SARS-CoV-2 infections in the district and lagged behind the MIS-C cases as shown *figure 3*.

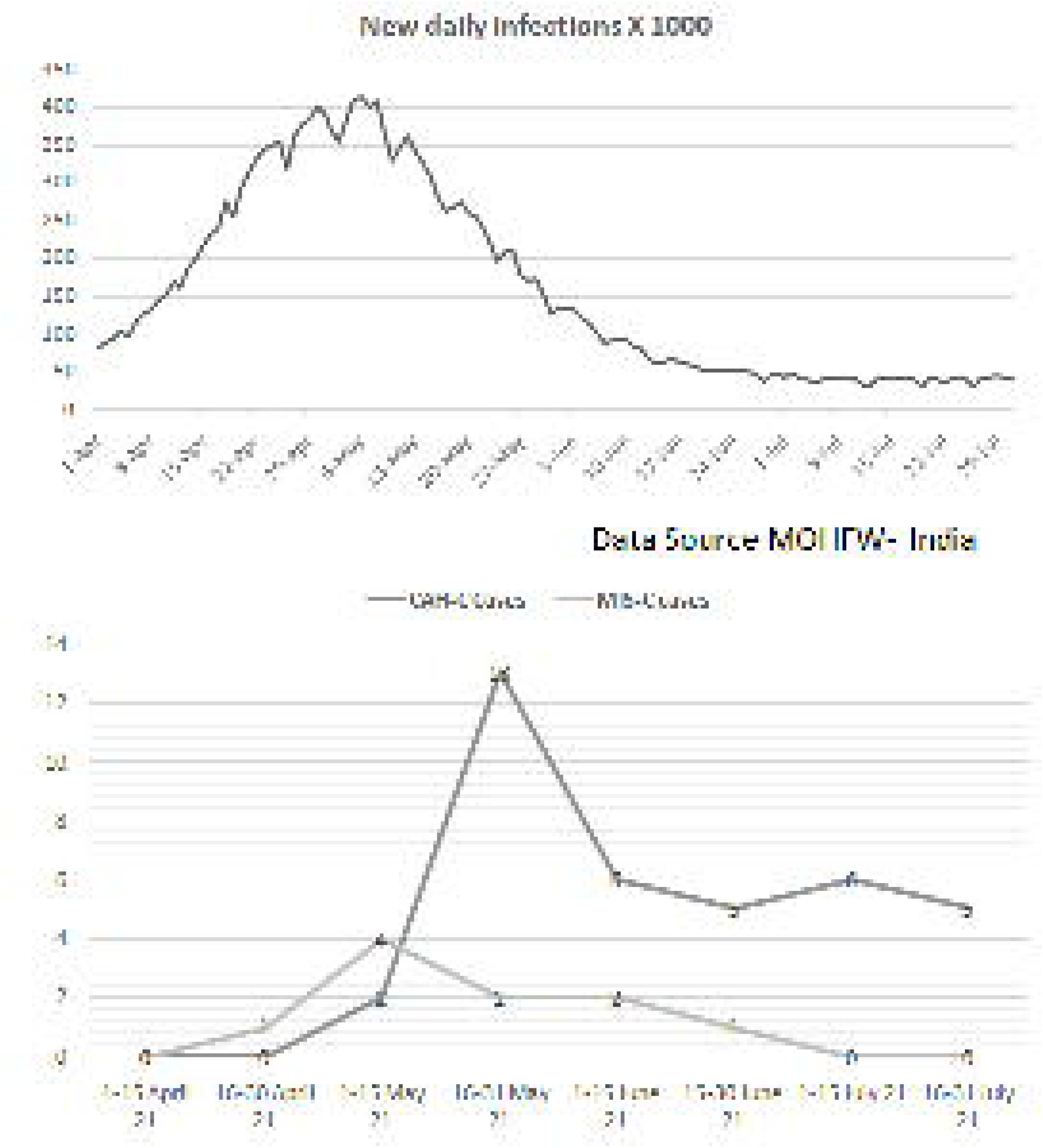

### Clinical and laboratory findings

CAH-C cases presented with typical symptoms of hepatitis including nausea, vomiting, and loss of appetite, weakness and mild fever not exceeding 38 º C. Two cases had signs of acute liver failure including altered sensorium. Ultrasound findings wherever available (n=5) were suggestive of periportal and cholecystic involvement, with GB wall thickening in one. None of them had any significant finding on chest X-ray (*figure 4)*.

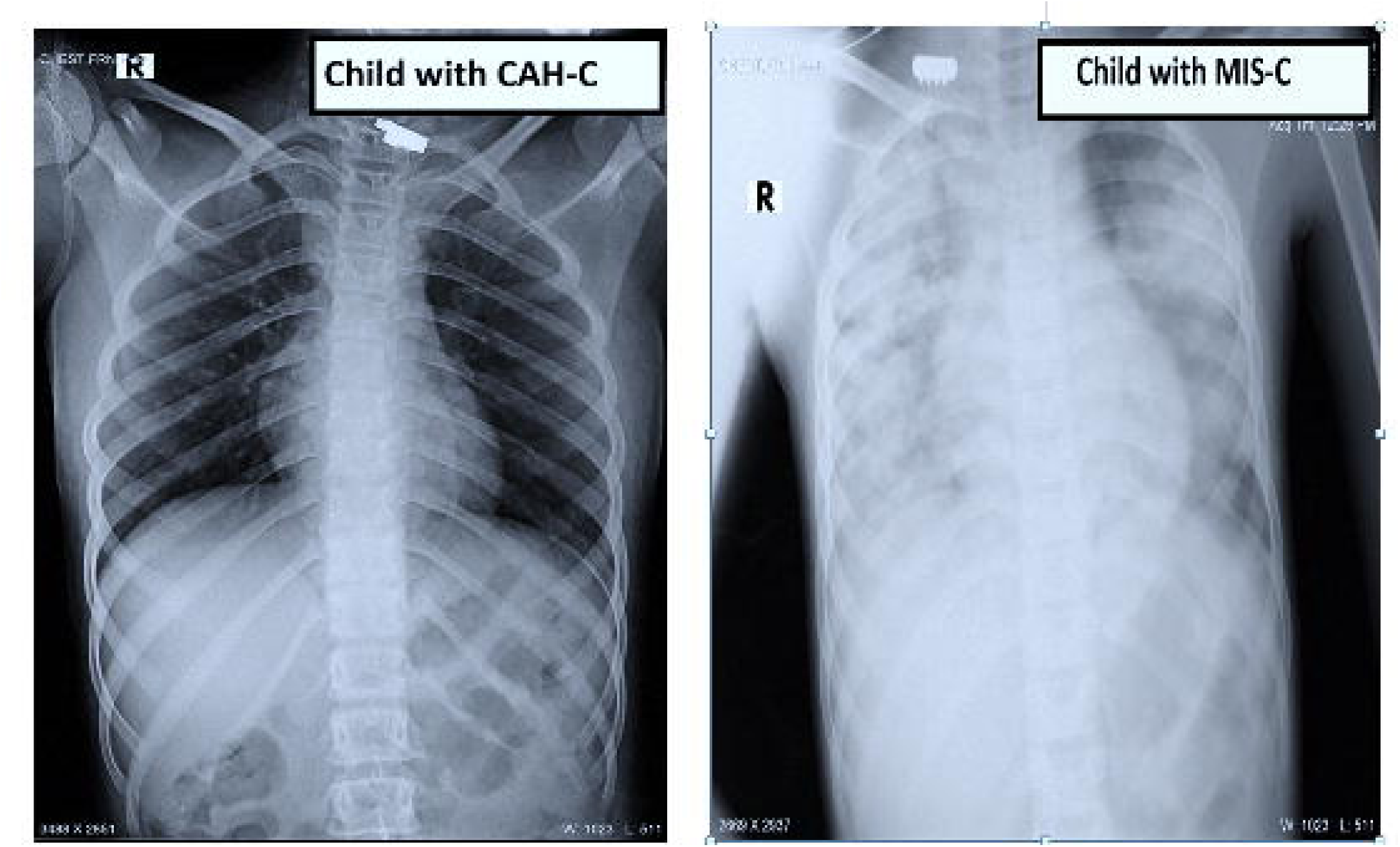

On laboratory investigation: 35/37 CAH-C cases had RTPCR test negative for SARS-CoV-2 by the time when admitted, majority cases (29/37) had significantly elevated transaminases (>10X upper limit of normal (ULN) with median 1326.2 (range 70-5685) U/L), serum bilirubin with median 4.05 (range 1.4-17.1) mg/dl, 25/37 had unelevated (remaining 12/37 had moderate elevation) C reactive protein (CRP), median 5.4 (range 0.70-7.9) mg/L, IL-6, median 9.7 (range 2.1-24.6.3) pg/ml (p=0.0001). Many (16/37) had elevated alkaline phosphatases 2X ULN (p= 0.29), slightly elevated total IgG levels median 1245.3 (range 551.4-1640) mg/dl. 35/37 had normal platelet counts, median 2.45 (range 0.69-7.7) /mm^3^×10^3^. These two children having thrombocytopenia also had signs of acute liver failure and had elevated prothrombin time and raised INR and D-Dimer. INR could be performed in 24/37 and D-Dimer in 8/37, among them 3 were raised and 5 were within range (*table 2*). All 37 (100%) patients were positive for SARS-CoV-2, anti-N protein antibodies in high titers. The results for tests performed for pathogens are shown in *table 3*.

**Table 2:**
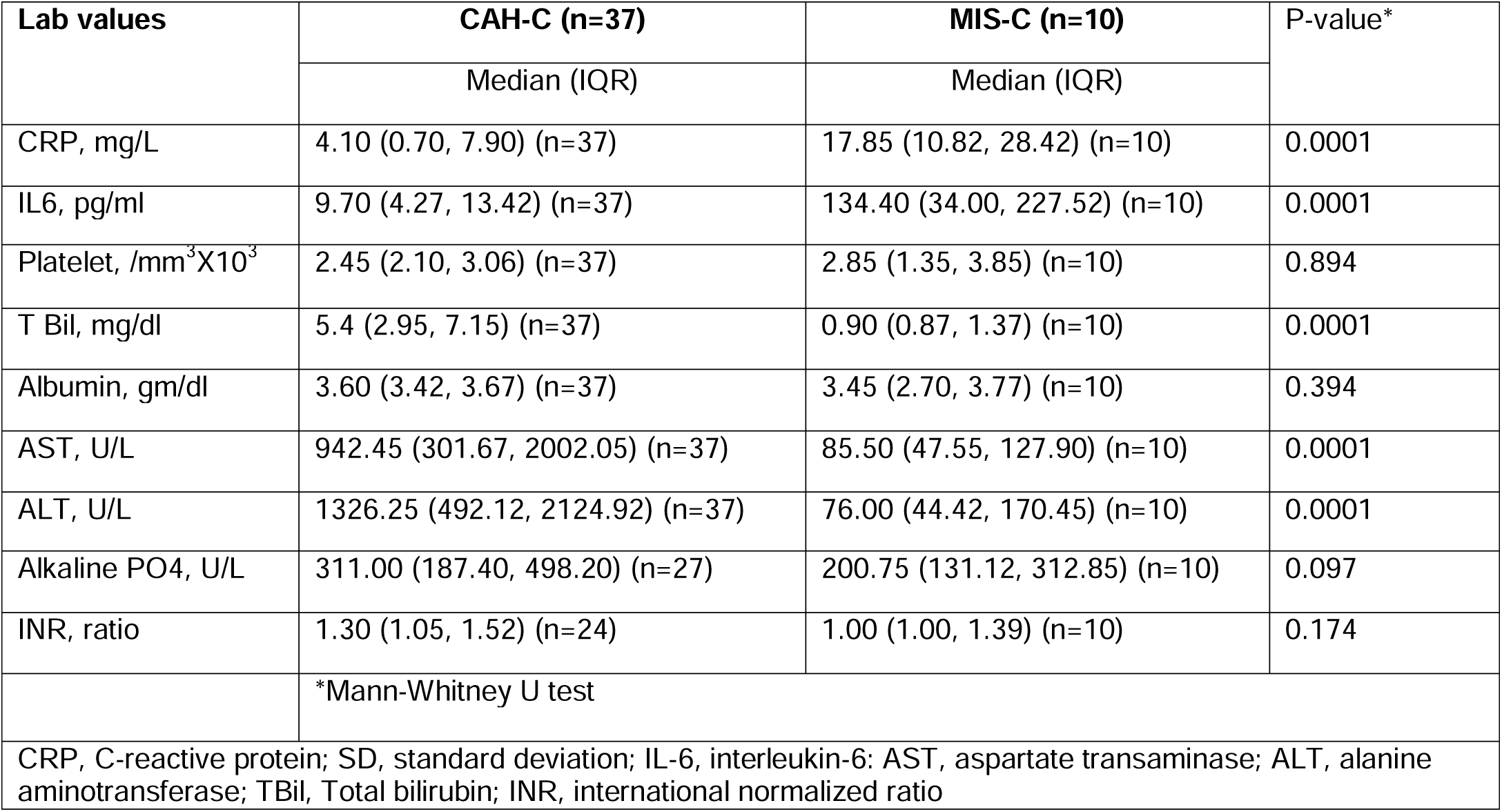
Laboratory findings of CAC-H Vs MIS-C associated hepatitis cases.

**Table 3:** Pathogens tested for and their results.

| Tests performed | CAH-C (total n=37) | MIS-C (total n=10) |
| --- | --- | --- |
|  | Positivity detected (% , tested n) | Positivity detected (% , tested n) |
| Anti- SARS-CoV-2 IgG (anti-N) | 37 (100%, tested n=37) | 3 (60%, tested n=5) |
| ANA | 0 (0%, tested, n=37) | 1 (tested, n=5) |
| Anti-LKM | 0 (0%, tested, n=37) | 0 (tested, n=5) |
| Anti-HAV IgM | 0 (0%, tested, n=37) | 1 (tested, n=8) |
| HbSAg | 0 (0%, tested, n=37) | 0 (0%, tested, n=8) |
| Anti-HCV | 0 (0%, tested, n=37) | 0 (0%, tested, n=8) |
| Anti-HEV IgM | 0 (0%, tested, n=37) | 0 (0%, tested, n=8) |
| Anti leptospira IgM | 0 (0%, tested, n=37) | 0 (0%, tested, n=5) |
| Anti VZV IgM | 7 (18.9%, tested n=37) | 0 (0%, tested, n=5) |
| Anti-EBV IgM | 8 (21.6%, tested n=37) | 0 (0%, tested, n=5) |
| Salmonella sp. (Widal) (titer>160) | 21 (56.7%, tested n=37) | 0 (0%, tested, n=8) |
| Dengue IgM | 8 (21.6%, tested n=37) | 0 (0%, tested, n=5) |
| HSV 1 & 2 (PCR) | 0 (0%, tested, n=17) | 0 (0%, tested, n=3) |
| Adenovirus (PCR) | 3 (17.6%, tested n=17) | 0 (0%, tested, n=3) |
| SARS- CoV-2 (PCR) | 34 (100%, tested, n=34) | 7 (70%, tested, n=10) |
| SARS-CoV-2 (repeat/on admission) | 2 (0.54%, tested n=37) | 3 (30%, tested, n=5) |

MIS-C with hepatitis cases presented with moderate to severe symptoms and a persistent fever of >38 ºC made them to be hospitalized. One child presented with epidermal necrosis and toxic shock like picture, one with encephalitis and 5 (50%) with pneumonia on chest X-rays (*figure 4)*. Besides persistent fever, they had sub-conjunctival hemorrhages, cough, shortness of breath, with features of hepatitis such as abdominal pain, loss of appetite and weakness. Of them, 3(30%) developed ARDS and signs of multi-organ failure mainly hypotension, one developed encephalitis and one developed signs of acute liver failure. All had admission in intensive care units.

On laboratory investigation (*table 2*): MIS-C patients presented with hepatitis, majority (7/10) were RT-PCR test positive for SARS-CoV-2 within 2-3 weeks, they had markedly raised CRP median 17.8 mg/L (p=0.0001), significantly elevated IL-6 median 134.4 pg/ml (p=0.0001) and reduced albumin levels with median 3.3 gm/dl (2.7-3.58) (p=0.116). D-Dimer level was elevated (>1.5) in all 7 cases in which it could be measured. They had reduced platelet counts in some cases, moderately elevated transaminases (ALT 40-200 U/L), normal or borderline raised serum bilirubin, mildly elevated alkaline phosphatases, and elevated procalcitonin (available for 4/10 cases). Anti SARS-CoV-2 IgG antibody test was positive in 3/5 cases; these 3 cases had a contact history with confirmed COVID-19 cases but were themselves RT-PCR negative.

Treatment (Total n=47): CAH-C cases (n=37), were admitted in non-critical care wards were given supportive therapy consisting of anti-emetics, IV fluids, vitamins, Zinc without any requirement of oxygen administration or steroids.

Those with MIS-C and hepatitis (n=10) were treated as per the ICMR recommended COVID-19 regimen for children, admission to critical care and other supportive treatment inclusive of IVIG (n=1) in child with neurologic symptoms, steroids in all, and oxygen administration (n=3) with mechanical ventilation in 1 case.

### Outcomes

All patients admitted with CAH-C were discharged with supportive treatment without any major complications with mean hospital stay of 4(± 1.0) days and remained uneventful at 4 weeks’ follow-up. No mortality was reported in this group.

Among the MIS-C children one child developed paralytic ileus which resolved on conservative treatment, however 3 out of 10 (30%) had an adverse outcome. The mean hospital stay in this group was 8(± 1) days. On 4 weeks’ post-discharge follow-up the remaining children had uneventful recovery.

## Discussion

With the emergence of newer VOC’S causing recurring waves of the pandemic, varied symptoms and post COVID-19 complications have been observed posing safety concerns even for the pediatric age group. In this scenario our study identified 37 cases with a unique presentation of acute hepatitis designated as CAH-C, whereas MIS-C could account for hepatitis in 10 cases amongst 15873 children screened in the district during the study period.

Cases of CAH-C suddenly presented at 2-6 weeks after remaining asymptomatic or mild symptomatic post-exposure to a lab-confirmed case of COVID-19. This might just be the tip of the iceberg, owing to the fact that this is an initial and early report of such cases. A possibility of missing some other less prominent features of this entity might be there which only time will tell, since initially we were focused mainly on the most prominent presenting symptoms. The spread of Delta or further variants of Concern in the anticipated 3^rd^ Phase of the pandemic warrants high alertness in detecting more of them.

CAH-C cases reported were greater in older children (6-11 years) and with male preponderance. Higher expression of receptors namely ACE2 in cholangiocytes**(23)** and TMPRSS2 in hepatocytes among males might be the predisposing factor**(24)**.

Prima facie, the findings of CAH-C cases do not resemble that of MIS-C, they lacked the markedly elevated inflammatory markers or systemic derangements seen in MIS-Cat all times of testing**(25)**. Interestingly, these cases had distinct immunological imprints having raised anti SARS-CoV-2 antibodies (titers >50 COI) in all the cases, significant Widal titers (>1:160 O, H), along with positive dengue IgM, positive anti-EBV IgM was seen in 8/37 cases and anti-VZV IgM in 7/37 patients respectively. The multi-positivity for various infectious agents along with elevated total IgG levels were missing in children having MIS-C and are unusual for children of such young age. These findings point to skewed immune activation, especially B cell stimulation giving rise to multiple antibody responses as a result of possible polyclonal B cell immune activation by some of the SARS-CoV-2 antigens or their possible cross reactions with these. This role played by SARS-CoV-2 in CAH-C is in contrast to the polyclonal (Vβ 21.3+ CD4+ and CD8+) T-cell stimulation observed in cases of MIS-C**(26)**. Thus, SARS-CoV-2 is a potent immune-stimulator through molecular mimicry leading to polyclonal B cell activation exclusively in children with a transient self-limiting course in CAH-C cases.

MIS-C being more severe form is hallmarked by polyclonal T-cell activation whereas CAH-C as observed is on the milder spectra of disease course hallmarked by polyclonal B-cell activation. Currently, these antibodies seem to be more of an evil, rather than a virtue offering any protection, since children exhibit narrow spectrum of reactivity against SARS-CoV-2 which may not be neutralizing in nature. Similar situation caused persistence of symptoms in adults recovered from COVID-19 due to alterations and persistence in B-cell responses during convalescence**(27)**. However, it would be premature to predict with certainty that CAH-C is a milder complication of the disease at this point. The other concern is the association of biological false positivity (BFP) which might pose a diagnostic dilemma in cases of other infectious diseases, wherein similar febrile illnesses including dengue, chikungunya and enteric fever remain endemic in developing countries. Both these facts warrant further elaborate studies amongst larger populations and also during the vaccine trails among children.

CAH-C cases were monitored very cautiously for deterioration since they were observed for the first time with no existing guidelines for treatment of such cases, none required specific treatment for COVID-19**(28)**. Interestingly they responded well following supportive treatment and care, most likely this high alertness prevented adversities among them. They had a shorter hospital stay, fair recovery, without any mortality at 4 weeks’ follow-up. A longer follow-up is desirable for more of these cases, along with elaborate experimental studies to develop appropriate treatment guidelines for this entity which was currently beyond scope.

In contrast to CAH-C, majority of MIS-C cases typically presented within 2-3 weeks in relation to SARS-CoV-2 infection with moderate to severe symptoms, their numbers peaked earlier (*figure 3*).They had markedly raised inflammatory markers (CRP/ ferritin, IL-6), elevated D-Dimer and decreased albumin levels in all cases as reported previously, besides testing negative for antiviral and other fever etiology.

MIS-C is already known to have higher mortality and a poor survival rate, hence despite best of efforts, it was associated with longer hospital stay and worse outcome having 30% mortality**(6)**.

To conclude, the continuous arrival of newer variants of SARS-COV-2 is often presenting with newer entities among different age groups. Our research highlights that, specifically amongst children with recent exposure to SARS-CoV-2 the timely recognition of such entities like CAH-C should enable the treating physician in prioritizing cases requiring admissions and initiating specific therapies. Thus, preventing adverse outcomes and reducing excessive burden on healthcare resources warranted during the pandemic.

## Data Availability

All data related to the manuscript is available with the author.

https://cvstatus.icmr.gov.in/

https://main.icmr.nic.in/sites/default/files/press_realease_files/ICMR_PR_IgG_Elisa_30052020.pdf

## Data sharing and materials

All data related to the study including the data dictionary is available with the authors and can be made available for common use. The findings of the study or any of its tables and figures can be cited or shared but with due credits to the authors. All figures submitted in the manuscript are original work created by the authors and not copyrighted by any third party.

## Acknowledgement

The authors thank and acknowledge Dr Rupesh Sahu for his support in suggesting right statistical tools and valuable feedback in completing the statistical analysis of the study. We are obliged by Dr Anirudh Singh and Dr Ashish K Vyas AIIMS Bhopal, India for their suggestions in improvement of the manuscript and for assistance in the plagiarism check. We are thankful to the Indian council of medical research (ICMR) for VRDL grant provided to set up the lab in 2018, however none of these funds have been directly used in the current study.

